# Validation of a Six-Item Multiple Sclerosis Impact Scale Short Form (MSIS-SF) for Efficient Assessment of Patient-Reported Physical Disturbance

**DOI:** 10.1101/2025.02.06.25321807

**Authors:** James F. Sumowski, Sarah Levy

**Affiliations:** Corinne Goldsmith Dickinson Center for Multiple Sclerosis, Department of Neurology, Icahn School of Medicine at Mount Sinai

## Abstract

Patient-reported outcomes importantly reflect the lived experiences of persons with multiple sclerosis (MS); however, given the broad array of MS symptoms, comprehensive evaluation can be burdensome, especially with lengthy questionnaires. The MS Impact Scale queries twenty different physical difficulties (MSIS-20). In 1150 patients with MS, evaluation of internal consistency indicated redundancy in the MSIS-20 (Cronbach’s alpha: 0.96), but optimal internal consistency (0.90) of a six-item short form (MSIS-SF; first six MSIS items). The MSIS-20 and MSIS-SF showed comparably strong associations with objective physical disability. The MSIS-SF provides a psychometrically robust, less burdensome, and time-efficient option for assessing patient-reported physical disturbance.

## INTRODUCTION

Patient-reported outcomes (PROs) reflect the day-to-day lived experience of persons with a medical condition; PROs are therefore essential components of clinical and research evaluations.(1) Persons with multiple sclerosis (MS) can experience a broad array of symptoms, including gait disturbance, other sensorimotor difficulties, fatigue, cognitive problems, mood changes, sleep disturbance, sexual dysfunction, and pain. A comprehensive evaluation can therefore include numerous questionnaires assessing these and other outcomes, which can be time-consuming and burdensome, especially if surveys are lengthy. Note also that lengthy surveys may lead to less thoughtful responses due to respondent fatigue, frustration, or boredom. The MS Impact Scale (MSIS-29) is a well-established PRO, with a physical subscale (MSIS-20) assessing twenty different physical complaints.(2) The MSIS-20 is typically used to estimate overall patient-reported physical dysfunction rather than specific symptoms. Using data from a large MS cohort, we aimed to develop a short form with equal or better psychometric properties.

## METHODS

The MS Center at Mount Sinai Hospital established a neurobehavioral screening clinic with the goal of monitoring functional outcomes for all patients with MS. A retrospective chart review approved by our Institutional Review Board (IRB) captured data from consecutive patients aged 18-65 years, diagnosed with MS, without another primary neurologic condition, serious mental illness (e.g., schizophrenia), or clinical relapse within two months. We also utilized data from an IRB-approved prospective research cohort enrolled using the same exclusion criteria, except with a more limited age range (20-50) and only relapsing-remitting disease course.(3)

### Short Form Internal Consistency

We evaluated internal consistency (Cronbach’s alpha) for the full MSIS-20 and then for progressively shortened forms by removing items one at a time, always removing the last remaining item. Items were removed in reverse order so that researchers could derive the short form from existing MSIS-20 data without risk of order effects. Ideal internal consistency (Cronbach’s alpha) is 0.90;(4) values >0.90 indicate redundancy (i.e., superfluous items). We therefore removed items until we reached ideal internal consistency (0.90), and then assessed the next two shorter iterations to confirm internal consistencies <0.90.

### Short Form Validity

We captured objective physical performance with the physical components of the MS Functional Composite (MSFC)(5) assessing walking speed (Timed 25 Foot Walk, T25FW(6)) and upper extremity coordination (Nine Hole Peg Test, NHPT(7)). Raw scores were converted to age- and sex-adjusted normative z-scores (NHPT using NIH Toolbox normative data; T25FW by adjusting for age and sex with general linear models, then using normative data(8)). Extreme z-scores (3.0*IQR) were winsorized to avoid undue influence of outliers before averaging z-scores into a composite physical performance metric. Rank-order correlations with 95% confidence intervals (95%CI) examined associations between composite physical performance and each MSIS form (MSIS-20, MSIS-SF). Pearson correlations were performed after assessment for skewness and kurtosis, and any necessary transformations. Norm-referenced z-scores were then used to categorize impaired performance (≤5^th^ percentile) on T25FW and NHPT; ANOVA compared differences in MSIS-20 and MSIS-SF (transformed if necessary) across patients with impairments on 0, 1, or 2 physical performance tasks. Differences in effect sizes (r, ρ, η^2^; 95%CIs) between MSIS-20 and MSIS-SF were compared.

## RESULTS

MSIS data were captured for 1150 persons with MS (mean [sd] age, 42.1 [11.4 years]; 71.2% [n=819] female; 83.9% [n=965] relapsing, 16.1% [n=185] progressive). As shown (Figure 1), internal consistency was high (indicative of redundancy) for the full MSIS-20 (0.963), but was closest to optimal (0.90) for a 6-item MSIS-SF (0.904) comprised of the first six MSIS-20 items.

**Figure 1.**
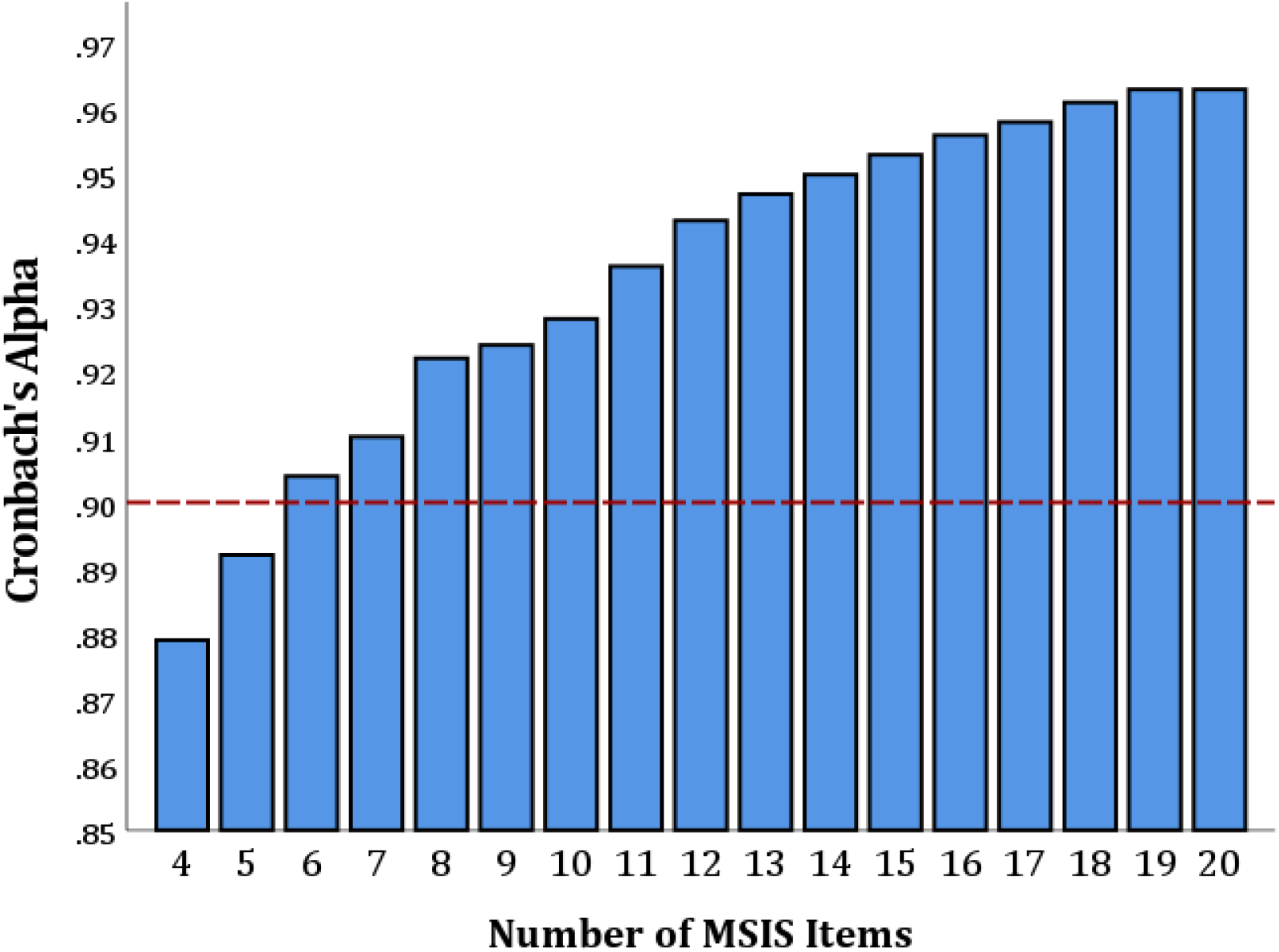
Internal Consistencies by Number of MSIS Items. Internal consistencies (Cronbach’s alpha) gradually decreased from 0.963 for the full 20-item MSIS until reaching the six-item short form (0.904) closest to optimal internal consistency (0.90, red dashed line); removal of additional items reduced internal consistency to below optimal.

Physical performance data (T25FW and NHPT) were available for 95.3% of the sample (n=1096); mean [sd] composite z-score, -0.62 [1.01]). Rank-order correlations (ρ [95%CI]) between physical performance and patient-reported physical difficulty were strong with no difference in effect sizes when using MSIS-SF (ρ= -0.523 [-0.566, -0.477]) versus MSIS-20 (ρ= -0.533 [-0.575, -0.488]). Mild skewness was observed for MSIS-SF (0.869), MSIS-20 (1.116), and physical performance (−1.005), which was rectified with log transformation of MSIS-SF and MSIS-20, and winsorization (1.5*IQR, n=48) of physical performance (all skewness <0.80).

Pearson correlations (r [95%CI]) similarly showed strong associations between physical performance and patient-reported physical difficulty, with no difference in effect sizes when using MSIS-SF (r= -0.564 [-0.603, -0.522]) versus MSIS-20 (r= -0.592 [-0.629, -0.552]). Likewise, ANOVA showed that patient-reported physical difficulty strongly differed across levels of impaired physical performance (Figure 2), with no difference in effect size when using MSIS-SF (η ^2^= 0.260 [0.217, 0.300]) versus MSIS-20 (η^2^= 0.284 [0.241, 0.324]).

**Figure 2.**
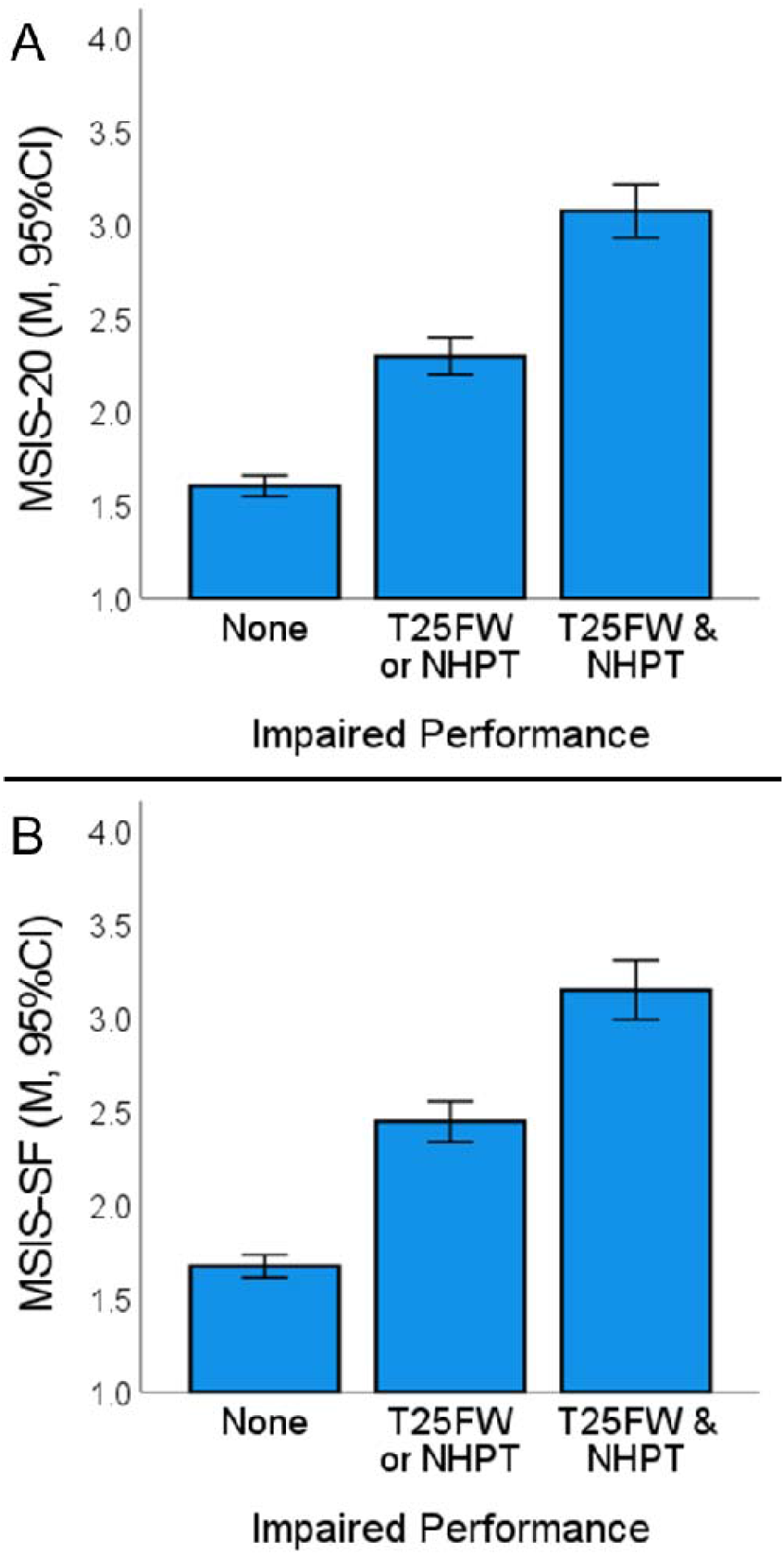
MSIS-20 and MSIS-SF across Levels of Objective Physical Impairment. Patient-reported physical difficulty assessed with (A) MSIS-20 (F[2, 1093]=216.55, p<0.001, η^2^ =0.284) and (B) MSIS-SF (F[2, 1093]=191.93, p<0.001, η^2^ =0.26) across level of physical impairment (None, n=754; T25FW or NHPT, n=232; T25FW & NHPT, n=110). All pairwise comparisons were significant at Bonferroni-adjusted P<0.001

Data were captured for an additional 293 patients (meeting the same selection criteria) who completed the six-item MSIS-SF after we replaced the MSIS-20 with this short form in our clinic (a change informed by the results presented herein). These patients did not differ in age (mean [sd], 41.6 [11.1]), sex [71.7% female], or disease course [12.6% progressive] from the sample of 1150 who completed the MSIS-20. MSIS-SF mean did not differ whether it was derived as the first six items of the full MSIS-20 (n=1150; mean [95%CI], 2.01 [1.95, 2.06]) versus when it was administered as the standalone six-item MSIS-SF (n=293; 2.01 [1.89, 2.13]).

## DISCUSSION

Findings support the six-item MSIS-SF as a psychometrically robust alternative to the full MSIS-20 for assessment of patient-reported physical difficulty. Analysis of internal consistency indicated redundancy in the full MSIS-20, which reduced to the optimal Cronbach’s alpha (0.90) in the MSIS-SF. Associations with objective physical performance were comparable for MSIS-20 and MSIS-SF. The full MSIS-20 may still make sense for clinicians or investigators interested in specific individual symptoms not queried by the MSIS-SF (e.g., spasticity, bladder symptoms), but the MSIS-SF provides a much more efficient option for clinicians and researchers interested in estimating overall patient-reported physical disturbance. We also demonstrated that MSIS-SF scores are the same whether derived from the first six items of the MSIS-20 or as the standalone six-item MSIS-SF, indicating that clinicians and researchers already using the MSIS-20 can confidently derive the MSIS-SF from existing data retrospectively and then switch to MSIS-SF henceforth. Validation of short forms of lengthy questionnaires for other symptoms is encouraged to optimize psychometric properties and reduce patient burden, as has been accomplished for cognition(9) and fatigue.(10)

## Data Availability

All data produced in the present study are available upon reasonable request to the authors.

